# Cardiovascular and Safety Events in Resistant Hypertension Treated with Transdermal Clonidine versus Spironolactone; A US Cohort Analysis in 33,753 individuals

**DOI:** 10.64898/2026.06.26.26356726

**Authors:** Nicole Princic, Megan Richards, Emmanouil Petrou, Claudio Borghi, George S Stergiou

## Abstract

**Objectives:** To compare real-world cardiovascular outcomes and safety events in patients with resistant hypertension following initiation of transdermal clonidine (TC) or spironolactone.

**Methods:** A retrospective analysis was performed using Merative™ MarketScan® Databases in the USA to identify cohorts with resistant hypertension initiating TC or spironolactone as a fourth-line agent between January 2012 and September 2024. Major Adverse Cardiovascular Events (MACE) and safety events were assessed during variable follow-up periods. Inverse probability of treatment weighting (IPTW) was applied to adjust for differences in baseline characteristics. Cox proportional hazard models were used to adjust for post-index beta-blocker utilization as a time-varying covariate for MACE outcomes.

**Results:** The analysis included 3,113 patients in the TC cohort and 30,640 in the spironolactone cohort. After IPTW, baseline characteristics were well balanced between cohorts (standardized mean differences <0.10; mean age 60 years, 54% male). Mean follow-up was 7.1 and 10.5 months for the TC and spironolactone cohorts, respectively. After IPTW no differences in MACE outcomes were observed between the two cohorts (weighted rate ratio 1.27 [0.79-2.06]). Results were consistent after adjusting for post-index beta-blocker use. The risk of hyperkalemia was significantly lower in the TC cohort (weighted rate ratio, 0.48 [0.33-0.70].

**Conclusions:** In this real-world analysis, patients with resistant hypertension treated with TC have similar risk for MACE outcomes as with spironolactone, but with significantly lower risk of hyperkalemia. Thus, in patients with resistant hypertension TC appears to provide similar cardiovascular protection, with a more favorable safety profile.

## INTRODUCTION

Hypertension is a major public health issue as it is the leading cause of premature death due to Major Adverse Cardiovascular Events (MACE) more than any other risk factor.^1–3^ Current guidelines recommend <130/80 mmHg as the optimal office blood pressure (BP) target with treatment, which often requires more than three first-line drug classes.^4, 5^ Resistant hypertension is defined as BP which remains uncontrolled despite pharmacologic treatment with angiotensin converting enzyme inhibitors (ACEi) or angiotensin receptor blockers (ARB), a calcium channel blocker (CCB), and a diuretic in maximum tolerated doses.^6–8^

Patients with resistant hypertension are at high risk of organ-damage and of cardiovascular and renal outcomes.^8–11^ The global prevalence of “apparent” resistant hypertension is estimated to be 10-20% of patients treated for high BP.^12, 13^ However, the prevalence of “true” resistant hypertension is lower (5-10%) and is challenging to diagnose, as a considerable proportion of “apparent resistance” is due to white coat hypertension, inadequate treatment regimen, poor medication adherence, etc.^14, 15^ Aldosterone antagonists (spironolactone) are recommended as the preferred fourth drug in patients resistant hypertension. Alternatively, any other antihypertensive drug not already utilized can be considered, including an a1-blocker (e.g., doxazosin), a centrally acting antiadrenergic drugs (e.g., clonidine), or a beta-blocker.^13,16–18^

The transdermal formulation of clonidine (TC), which is available in the US and in Italy, is a potentially underutilized treatment option for resistant hypertension.^19^ TC is an interesting pharmacotechnical formulation of clonidine, with BP lowering efficacy comparable to first-line antihypertensive agents^20–22^, a unique once-per-week administration which improves adherence with treatment, and considerably fewer adverse effects than oral clonidine.^20–22^ This study investigated whether TC is a useful alternative to spironolactone for patients with resistant hypertension in terms of cardiovascular protection and adverse effects.

## METHODS

### Study Design and Data Source

This is a retrospective longitudinal cohort study conducted using claims from the USA Merative™ MarketScan® Commercial and Medicare Databases^23^ between 1 January 2012 and 30 September 2024 (“study period”). These databases include beneficiaries and their dependents covered under a variety of fee-for-service and managed healthcare plans.

Variables were defined using International Classification of Diseases, 9^th^ and 10^th^ Revision, Clinical Modification (ICD-10-CM) codes, Current Procedural Terminology (CPT) 4th edition codes, Healthcare Common Procedure Coding System (HCPCS) codes, and National Drug Codes (NDC). All database records are de-identified and certified to be fully compliant with the US patient confidentiality requirements set forth in the Health Insurance Portability and Accountability Act of 1996. As this study uses only de-identified patient records and does not involve the collection, use, or transmittal of individually identifiable data, the data does not involve human subjects (per the definition of human subjects in the Code of Federal Regulations [CFR] Title 45 Part 46.102[e]). Thus, an Institutional Review Board [IRB] approval for performing the present analyses is not required.

### Patient Selection and Cohort Development

Patients with resistant hypertension were identified between 1 January 2012 and 30 September 2024 using criteria consistent with guidelines as applied in prior real-world studies, defined as those with uncontrolled office BP while on concomitant use of three antihypertensive agents including an ACE/ARB, a CCB and a thiazide or thiazide-like diuretic.^6, 8, 16, 17^ Episodes of concomitant use were determined using prescription fill dates and days of supply from each claim and defined as a period of overlapping days for all three classes.^6, 24^ Patients with resistant hypertension were selected for the two mutually exclusive TC or spironolactone cohorts if they newly initiated the treatment as a fourth drug as an add-on therapy while on the triple-drug regimen. The first prescription for TC or spironolactone was set as the index date.

Both cohorts were required to have at least 12 months of continuous enrollment with medical and pharmacy coverage prior to index (“baseline period”), at least one claim with a diagnosis of essential or primary hypertension during baseline, and be aged 18 years or older as of the index date. To ensure patients were newly initiating TC or spironolactone and that cohorts were mutually exclusive, patients were excluded if they had the index treatment during the baseline period and if they had the comparator drug during the baseline period or on the index date. In addition, patients were excluded if they had a diagnosis of primary aldosteronism or heart failure at baseline (see Table S1 for ICD-9-CM and ICD-10-CM diagnosis codes). Finally, patients with prior diagnoses of any of the cardiovascular outcomes or safety events at baseline were excluded. Although this exclusion had the potential to introduce selection bias it was necessary to ensure accurate measurement of incidence.

### Cardiovascular Outcomes and Safety Events

All cardiovascular outcomes and safety events were identified during a variable length follow-up period starting from the index date until evidence of an outcome, discontinuation of the index medication (defined as a 14 day gap following the end of drug supply), switch or addition of the comparator drug, end of database enrollment, or end of study period (30 September 2024), whichever event occurred first. To ensure results would not be subject to immortal time bias, no minimum follow-up duration was imposed.

The cardiovascular outcomes were MACE, including ischemic stroke and acute myocardial infarction. Ischemic stroke was defined by at least one diagnosis code for the condition as the primary discharge diagnosis on inpatient claims, and acute myocardial infarction using inpatient claims with at least one diagnosis code for the condition in the primary or secondary position.^6, 25, 26^

The safety events included documented adverse events associated with index treatments and included constipation, diarrhea, dizziness/drowsiness, dry mouth, hyperkalemia, kidney function deterioration, nausea/vomiting, and skin rash. Kidney function deterioration was defined using the following; at least one inpatient claim with a diagnosis of advanced chronic kidney disease (CKD) stage 3-5, or evidence of end-stage renal disease (ESRD) defined by at least two claims with ESRD or dialysis codes at least 30 days apart, or kidney transplant.^27^ CKD stage 3-5 or ESRD (and not earlier stage CKD) was selected to ensure capture of kidney disease progression or deterioration. A broader definition of CKD inclusive of stage 1-2, would capture new onset of kidney disease, but this would not necessarily be indicative of worsening disease and therefore was not included in the definition. All other safety outcomes were defined by at least one medical claim with a diagnosis code for the condition (see Table S2 for ICD-9-CM and ICD-10-CM diagnosis codes).

### Covariates

Demographic data, clinical characteristics, medications, healthcare utilization and costs were measured on index or at baseline, as these covariates could impact or be associated with selection of TC or spironolactone. Demographics included age, sex, USA Census Bureau geographic region, insurance plan type, payer, and index year. General health status was measured using the Charlson Comorbidity Index (CCI). The CCI is an aggregate measure of comorbidity expressed as a numeric score based on the presence of selected diagnoses for various conditions, each with specific weights ranging from 1 to 6 points.^28, 29^

Individual clinical conditions and drug classes measured at baseline are listed in Table 1 and Table S3. In addition, oral clonidine use was evaluated at baseline as patients may have utilized it prior to initiating the TC form. It was also essential to ensure patients with prior oral clonidine were included, to ensure a representative sample of all patients newly initiating the TC form which has a different adherence and adverse effect profile than the oral formulation.^21, 30^ Lastly, because beta-blockers are widely used in the management of various cardiovascular conditions including hypertension^5,31^ and therefore comprise a potential confounder particularly for MACE outcomes, their utilization was captured during both baseline and follow-up. The total number of prescriptions and proportion of days covered (PDC) by beta-blockers were also assessed.

**Table 1.**
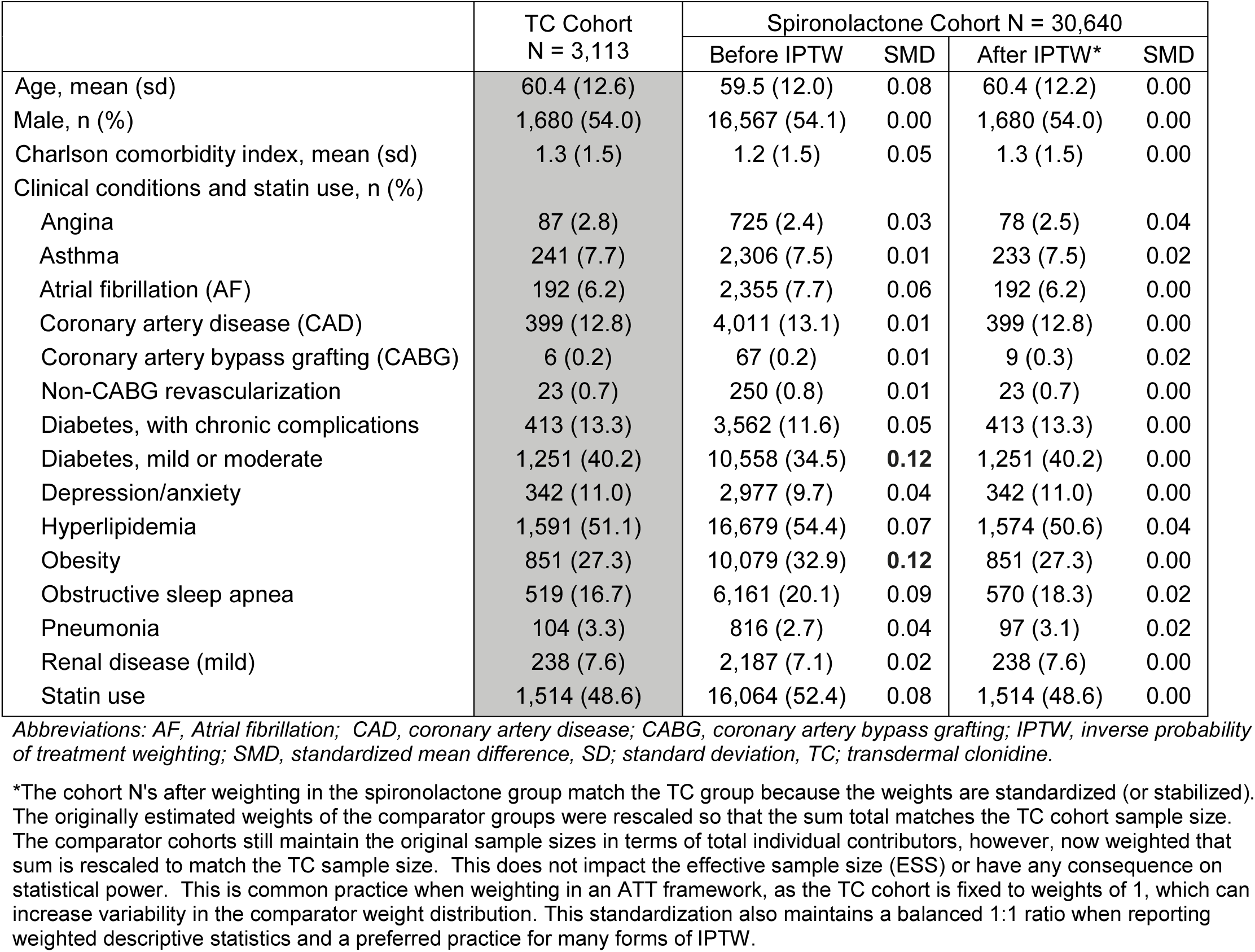
Baseline characteristics of patients with resistant hypertension newly initiating transdermal clonidine or spironolactone before and after inverse probability of treatment weighting.

|  | TC Cohort<br>N = 3,113 | Spironolactone Cohort N = 30,640 |  |  |  |
| --- | --- | --- | --- | --- | --- |
|  |  | Before IPTW | SMD | After IPTW* | SMD |
| Age, mean (sd) | 60.4 (12.6) | 59.5 (12.0) | 0.08 | 60.4 (12.2) | 0.00 |
| Male, n (%) | 1,680 (54.0) | 16,567 (54.1) | 0.00 | 1,680 (54.0) | 0.00 |
| Charlson comorbidity index, mean (sd) | 1.3 (1.5) | 1.2 (1.5) | 0.05 | 1.3 (1.5) | 0.00 |
| Clinical conditions and statin use, n (%) |  |  |  |  |  |
| Angina | 87 (2.8) | 725 (2.4) | 0.03 | 78 (2.5) | 0.04 |
| Asthma | 241 (7.7) | 2,306 (7.5) | 0.01 | 233 (7.5) | 0.02 |
| Atrial fibrillation (AF) | 192 (6.2) | 2,355 (7.7) | 0.06 | 192 (6.2) | 0.00 |
| Coronary artery disease (CAD) | 399 (12.8) | 4,011 (13.1) | 0.01 | 399 (12.8) | 0.00 |
| Coronary artery bypass grafting (CABG) | 6 (0.2) | 67 (0.2) | 0.01 | 9 (0.3) | 0.02 |
| Non-CABG revascularization | 23 (0.7) | 250 (0.8) | 0.01 | 23 (0.7) | 0.00 |
| Diabetes, with chronic complications | 413 (13.3) | 3,562 (11.6) | 0.05 | 413 (13.3) | 0.00 |
| Diabetes, mild or moderate | 1,251 (40.2) | 10,558 (34.5) | **0.12** | 1,251 (40.2) | 0.00 |
| Depression/anxiety | 342 (11.0) | 2,977 (9.7) | 0.04 | 342 (11.0) | 0.00 |
| Hyperlipidemia | 1,591 (51.1) | 16,679 (54.4) | 0.07 | 1,574 (50.6) | 0.04 |
| Obesity | 851 (27.3) | 10,079 (32.9) | **0.12** | 851 (27.3) | 0.00 |
| Obstructive sleep apnea | 519 (16.7) | 6,161 (20.1) | 0.09 | 570 (18.3) | 0.02 |
| Pneumonia | 104 (3.3) | 816 (2.7) | 0.04 | 97 (3.1) | 0.02 |
| Renal disease (mild) | 238 (7.6) | 2,187 (7.1) | 0.02 | 238 (7.6) | 0.00 |
| Statin use | 1,514 (48.6) | 16,064 (52.4) | 0.08 | 1,514 (48.6) | 0.00 |

Abbreviations: AF, Atrial fibrillation; CAD, coronary artery disease; CABG, coronary artery bypass grafting; IPTW, inverse probability of treatment weighting; SMD, standardized mean difference; SD, standard deviation, TC; transdermal clonidine.
\*The cohort N's after weighting in the spironolactone group match the TC group because the weights are standardized (or stabilized). The originally estimated weights of the comparator groups were rescaled so that the sum total matches the TC cohort sample size. The comparator cohorts still maintain the original sample sizes in terms of total individual contributors, however, now weighted that sum is rescaled to match the TC sample size. This does not impact the effective sample size (ESS) or have any consequence on statistical power. This is common practice when weighting in an ATT framework, as the TC cohort is fixed to weights of 1, which can increase variability in the comparator weight distribution. This standardization also maintains a balanced 1:1 ratio when reporting weighted descriptive statistics and a preferred practice for many forms of IPTW.

Baseline healthcare utilization and costs were reported separately for inpatient services, outpatient services, and pharmacy prescriptions. Healthcare costs were based on paid amounts of adjudicated claims, including insurer and health plan payments, as well as patient cost-sharing in the form of copayment, deductible, and coinsurance. All US dollar estimates were inflated to 2024 dollars using the Medical Care Component of the Consumer Price Index (CPI).^32^

### Statistical analysis

All the baseline study measures were summarized descriptively. Categorical variables were presented as the count and percent of patients in each category. Continuous variables were summarized with means and standard deviations (SDs).

To minimize the potential treatment selection bias, inverse probability of treatment weighting (IPTW) in the average treatment effect on treated (ATT) framework was used to adjust for differences in baseline characteristics (Figure S1) between the TC and the spironolactone cohort. IPTW leverages propensity scores to weigh each patient according to the inverse of the probability of receiving the treatment received, thereby creating a pseudo-population in which the distributions of baseline patient characteristics emulated the TC cohort and are balanced between comparator treatment cohorts. Performance of the IPTW was accessed by summarizing baseline covariates before and after weighting and considered well balanced if the standardized mean difference (SMD) between cohorts was <0.1.^33,34^

Incidence rates were reported per 1,000 person years at risk. In addition, 95% confidence intervals and incidence rate ratios were reported after adjusting for differences in patient characteristics through IPTW. Kaplan-Meier curves were used to measure cumulative incidence of each cardiovascular outcome and safety event after IPTW. Plots containing the observed probability of each event comparing the TC vs. spironolactone were produced, with significance tested using Log-rank test. An alpha of <0.05 was considered as statistically significant.

Beta-blocker use can vary in the follow-up period and can confound the association of TC treatment with MACE. Thus, beta-blocker use was incorporated as a time-varying covariate in a Cox proportional hazard model using the counting process formulation. This approach partitions an individual’s follow-up data into intervals where beta-blocker use is constant, allowing beta-blocker history to update to new changes. Each segment is weighted using the IPTW previously constructed, and estimation was performed using partial likelihood.^35, 36^ Standard errors for each adjusted hazard ratio (aHR) were estimated using Huber-White robust standard error estimation, as the weights and partitions induce heteroskedasticity and intra-patient correlation. A visual summary of all study methods is provided in Figure S1.

## RESULTS

Among 2,522,648 patients identified as having apparent resistant hypertension, there were 16,206 and 106,306 patients initiating TC and spironolactone respectively, assuming a diagnosis of true resistant hypertension by healthcare professionals. After applying all study inclusion and exclusion criteria a total of 3,113 patients in the final TC cohort and 30,640 in the spironolactone cohort were included (Figure 1).

**Figure 1.**
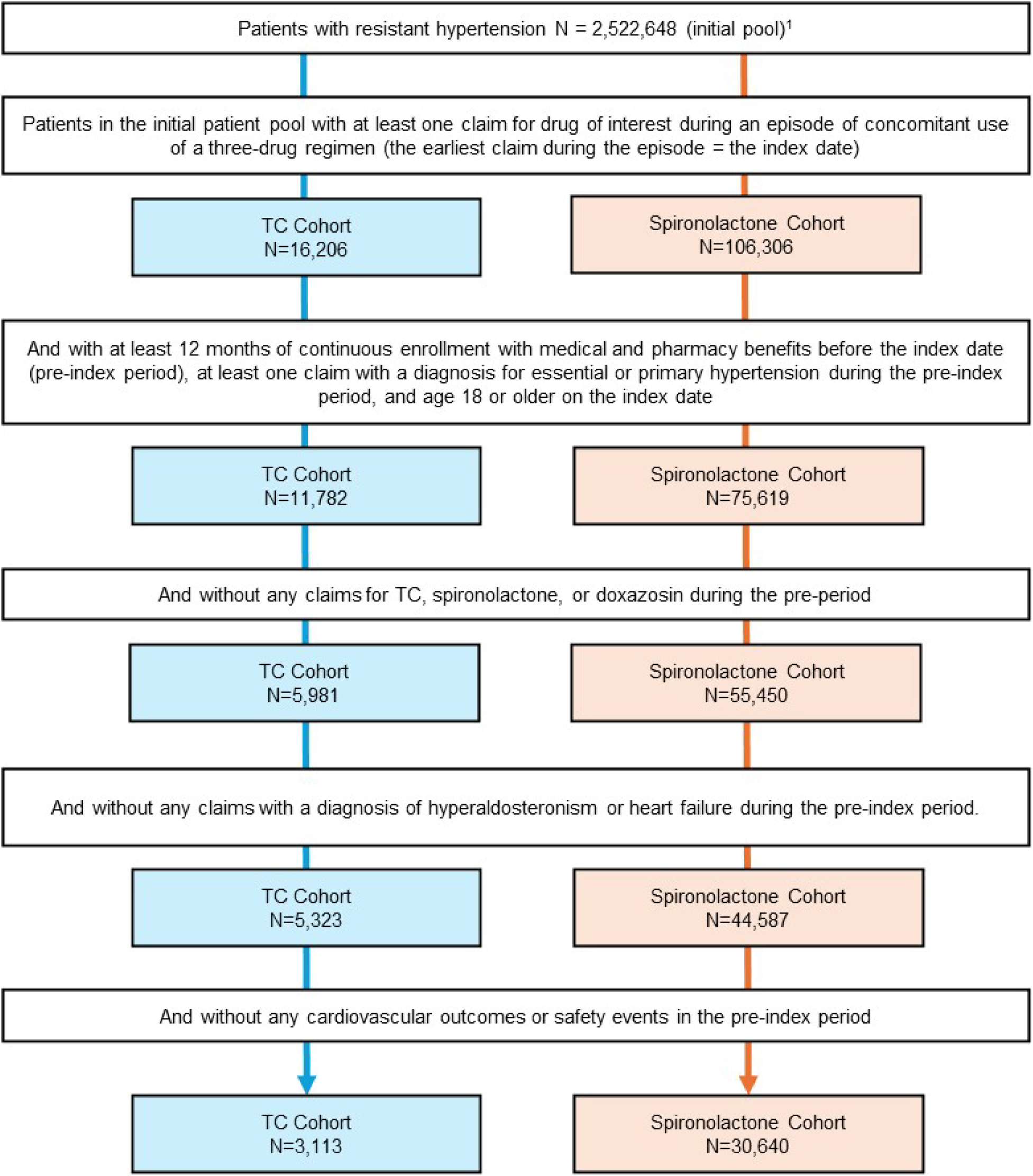
Patient selection. ^1^Eligible for the TC cohort if they met criteria for resistant hypertension; eligible for the spironolactone cohort if they met criteria for resistant hypertension and were not eligible for the TC cohort. *Abbreviations: TC; transdermal clonidine*.

Table 1 and Table S3 includes the baseline demographic, clinical, and treatment characteristics in the TC and the spironolactone cohort before and after IPTW. After weighting, baseline characteristics were well balanced (SMD <0.10 with most <0.05), with mean age of both study cohorts at 60 years, 54% males and index year distribution being identical in the two cohorts. Hyperlipidemia, diabetes, and obesity were the most common baseline comorbidities (51%, 40% and 27% respectively) and consistent with that finding, statins and antidiabetics were the most common non-antihypertensive treatments prescribed at baseline (mean of 2.3 and 2.7 fills respectively). After weighting, 61% of patients in both cohorts had at least one prescription for beta-blockers at baseline (29% in the immediate 30 days preceding index and 51% in the 90 days preceding index) and with an average of 3.1-3.2 prescriptions during the 12-month period (all with SMD <0.05).

Twenty-nine per cent of patients in the TC cohort had at least one prescription for oral clonidine during baseline, compared with 11% of the spironolactone cohort. This difference was expected as patients receiving oral clonidine may transition to the TC formulation. The average follow-up on the index treatment was 7.1 (SD 12.2) months for the TC cohort and 10.5 months (SD 15.4) for the spironolactone cohort, with a discontinuation gap (period of >14 days following the end of drug supply) being the most common event triggering the end of follow-up (71%-74%) in both cohorts, likely due to patient behavioral patterns. Incidence rates for each event were reported per 1,000 person years at risk to adjust for varying follow-up duration by using the aggregate time-at-risk as the denominator.

### Cardiovascular Outcomes

During follow-up, incidence of MACE was low in both the TC (n=19) and in the spironolactone cohort (n=22) with incidence rates of 10.6 and 8.3 events per 1000 person years at risk. Incidence rates were lower for the individual events including ischemic stroke (5.0 and 3.3 events per 1000 person-years) and acute myocardial infarction (5.6 and 5.2 events per 1000 person-years) for both TC and spironolactone cohorts respectively. The rate ratio after adjusting for differences in baseline characteristics using IPTW was numerically higher for the TC cohort (versus spironolactone) but not statistically significant either for MACE overall or for stroke or acute myocardial infarction (Table 2). Using COX proportional hazard models to adjust for differences in beta-blocker utilization during the follow-up period as a time-varying covariate (a potential confounder for MACE outcomes) reinforced the finding that there were no consistent statistically significant differences between groups (aHR 1.19 [95% CI: 0.74-1.92; p=0.48]). The cumulative incidence plots using Kaplan Meier plots after IPTW for all cardiovascular outcomes are provided in Figure 2A.

**Figure 2.**
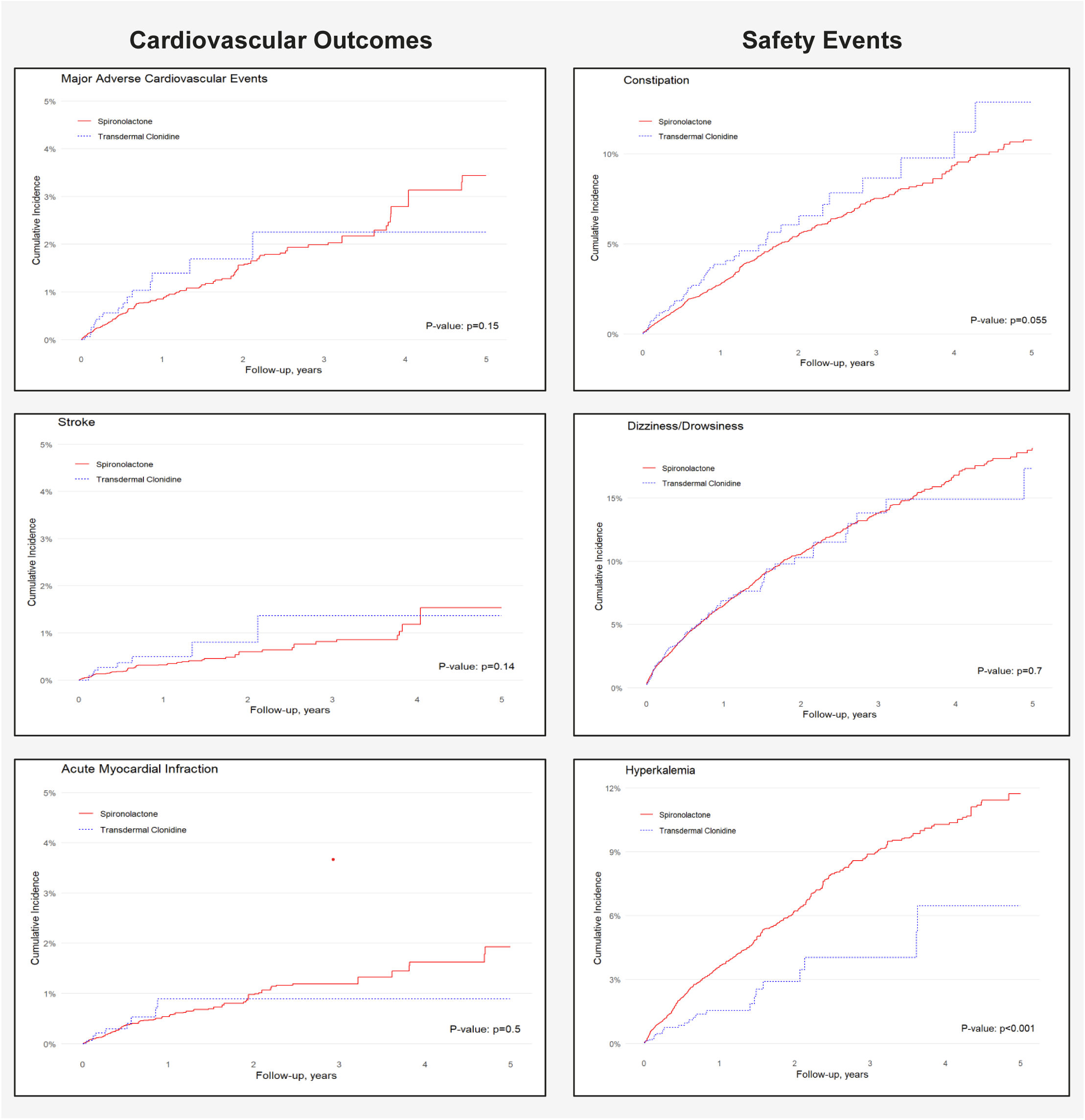
Cumulative incidence rates adjusted using inverse probability of treatment weighting among patients with resistant hypertension newly initiating transdermal clonidine versus spironolactone.

**Table 2.**
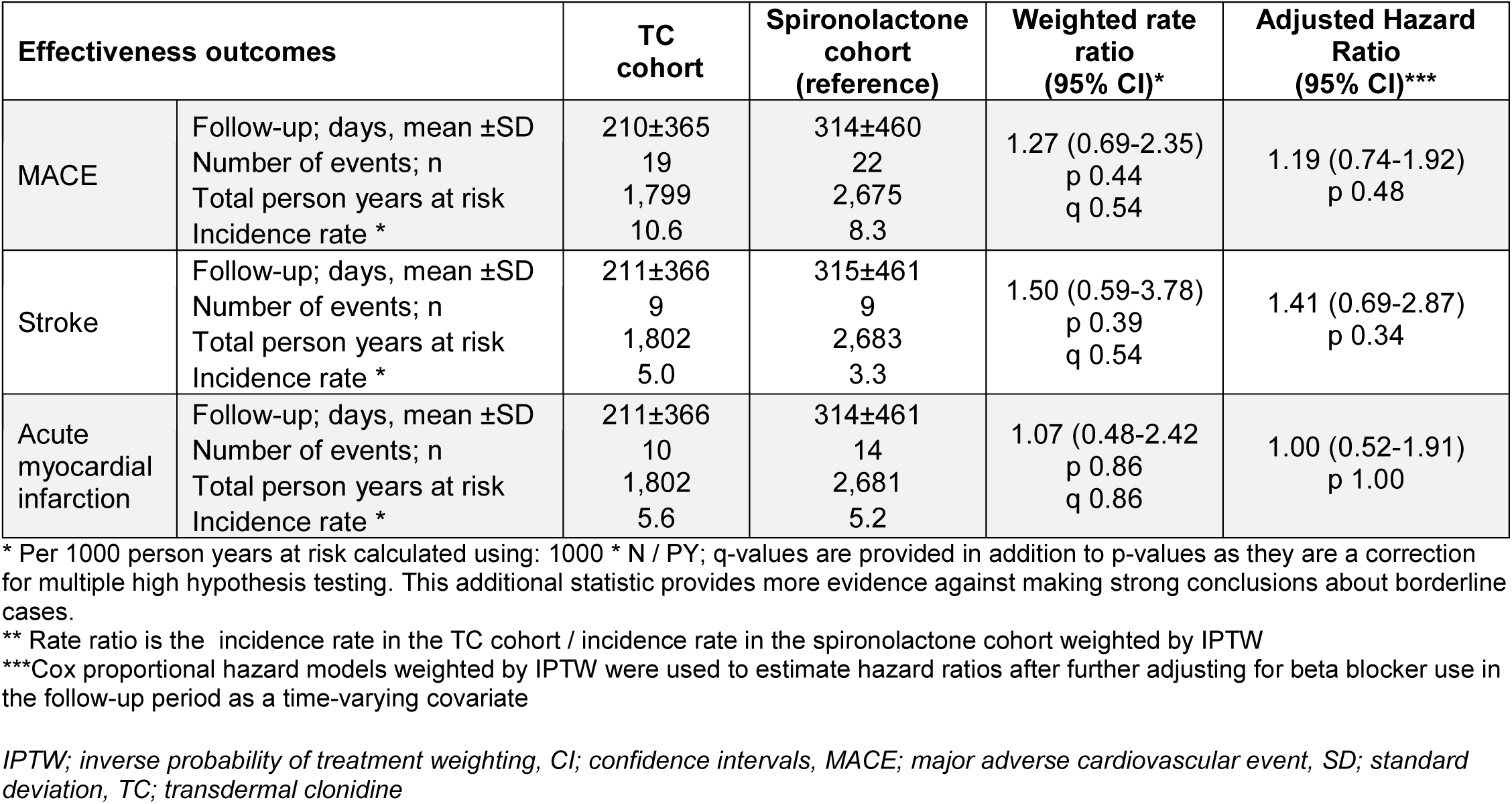
Cardiovascular outcomes – transdermal clonidine versus spironolactone; adjusted using inverse probability of treatment weighting.

### Safety Events

During follow-up, after adjustment for differences in baseline characteristics using IPTW, patients with TC had significantly lower risk of hyperkalemia compared with spironolactone (rate ratio 0.48 [95% CI: 0.32-0.73; p<0001) with an incidence rate of 16.3 versus 33.6 per 1000 person years at risk (Table 3), which was a highly statistically significant difference in cumulative incidence over time (p<0.001) with an early (within the first month) divergence between groups immediately following treatment initiation (Figure 2B).

**Table 3.**
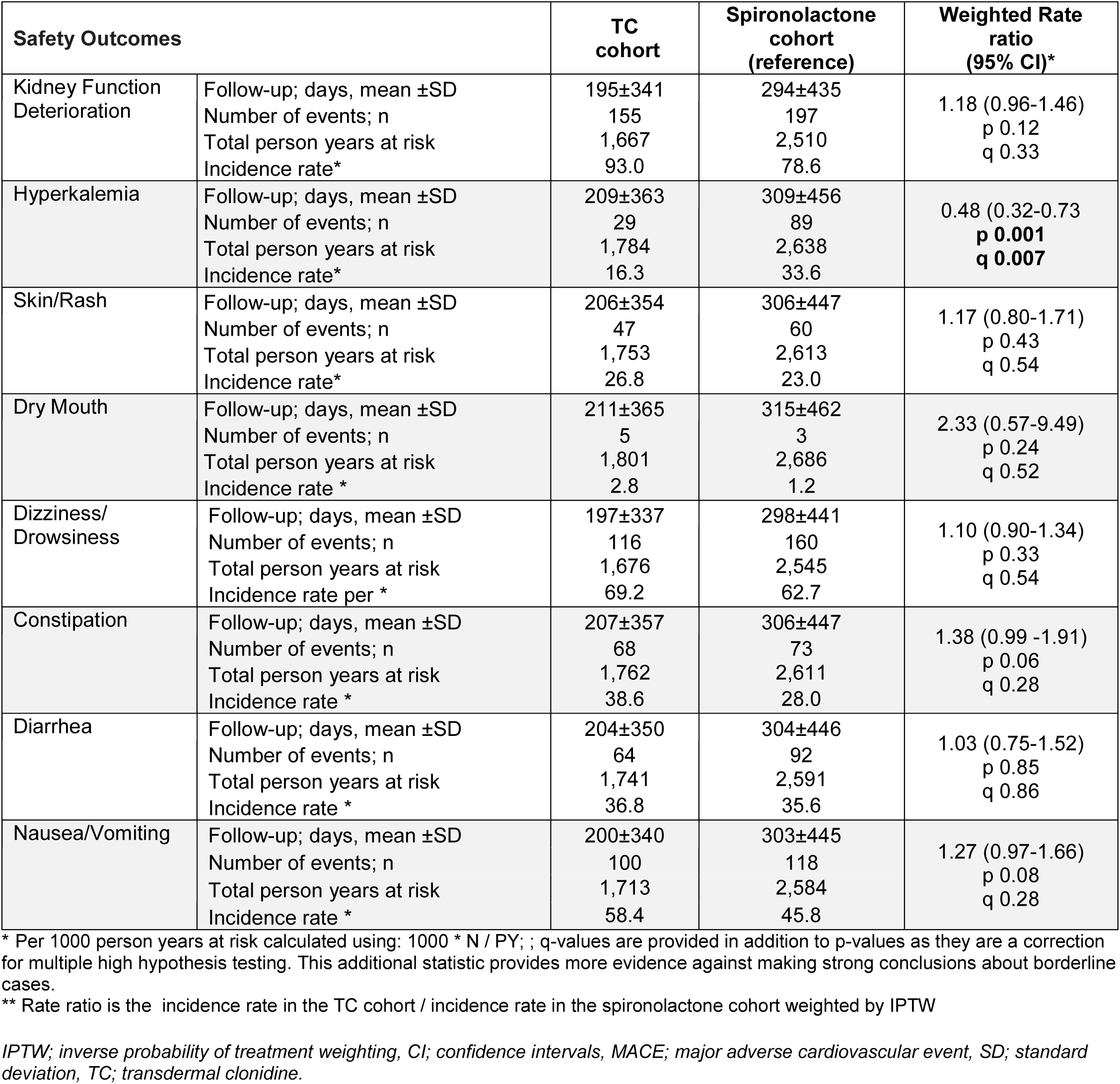
Safety events – transdermal clonidine versus spironolactone; adjusted using inverse probability of treatment weighting.

| Safety Outcomes |  | TC cohort | Spironolactone cohort (reference) | Weighted Rate ratio (95% CI)* |
| --- | --- | --- | --- | --- |
| Kidney Function Deterioration | Follow-up; days, mean ±SD | 195±341 | 294±435 | 1.18 (0.96-1.46)<br>p 0.12<br>q 0.33 |
|  | Number of events; n | 155 | 197 |  |
|  | Total person years at risk | 1,667 | 2,510 |  |
|  | Incidence rate* | 93.0 | 78.6 |  |
| Hyperkalemia | Follow-up; days, mean ±SD | 209±363 | 309±456 | 0.48 (0.32-0.73)<br>**p 0.001**<br>**q 0.007** |
|  | Number of events; n | 29 | 89 |  |
|  | Total person years at risk | 1,784 | 2,638 |  |
|  | Incidence rate* | 16.3 | 33.6 |  |
| Skin/Rash | Follow-up; days, mean ±SD | 206±354 | 306±447 | 1.17 (0.80-1.71)<br>p 0.43<br>q 0.54 |
|  | Number of events; n | 47 | 60 |  |
|  | Total person years at risk | 1,753 | 2,613 |  |
|  | Incidence rate* | 26.8 | 23.0 |  |
| Dry Mouth | Follow-up; days, mean ±SD | 211±365 | 315±462 | 2.33 (0.57-9.49)<br>p 0.24<br>q 0.52 |
|  | Number of events; n | 5 | 3 |  |
|  | Total person years at risk | 1,801 | 2,686 |  |
|  | Incidence rate * | 2.8 | 1.2 |  |
| Dizziness/<br>Drowsiness | Follow-up; days, mean ±SD | 197±337 | 298±441 | 1.10 (0.90-1.34)<br>p 0.33<br>q 0.54 |
|  | Number of events; n | 116 | 160 |  |
|  | Total person years at risk | 1,676 | 2,545 |  |
|  | Incidence rate per * | 69.2 | 62.7 |  |
| Constipation | Follow-up; days, mean ±SD | 207±357 | 306±447 | 1.38 (0.99 -1.91)<br>p 0.06<br>q 0.28 |
|  | Number of events; n | 68 | 73 |  |
|  | Total person years at risk | 1,762 | 2,611 |  |
|  | Incidence rate * | 38.6 | 28.0 |  |
| Diarrhea | Follow-up; days, mean ±SD | 204±350 | 304±446 | 1.03 (0.75-1.52)<br>p 0.85<br>q 0.86 |
|  | Number of events; n | 64 | 92 |  |
|  | Total person years at risk | 1,741 | 2,591 |  |
|  | Incidence rate * | 36.8 | 35.6 |  |
| Nausea/Vomiting | Follow-up; days, mean ±SD | 200±340 | 303±445 | 1.27 (0.97-1.66)<br>p 0.08<br>q 0.28 |
|  | Number of events; n | 100 | 118 |  |
|  | Total person years at risk | 1,713 | 2,584 |  |
|  | Incidence rate * | 58.4 | 45.8 |  |

\* Per 1000 person years at risk calculated using: $1000 * N / PY$ ; ; q-values are provided in addition to p-values as they are a correction for multiple high hypothesis testing. This additional statistic provides more evidence against making strong conclusions about borderline cases.
\*\* Rate ratio is the incidence rate in the TC cohort / incidence rate in the spironolactone cohort weighted by IPTW
*IPTW; inverse probability of treatment weighting, CI; confidence intervals, MACE; major adverse cardiovascular event, SD; standard deviation, TC; transdermal clonidine.*

Kidney function deterioration was the safety event that had the highest incidence in both cohorts (93.0 and 78.6 events per 1000 person-years at risk for the TC and spironolactone cohorts respectively) with a rate ratio of 1.18 (95% CI: 0.96-1.46; p=0.12) (Table 3). The cumulative incidence of kidney function deterioration was also higher with TC compared to spironolactone (p=0.04), however although baseline kidney disease diagnoses were adjusted for, some residual imbalance may remain because laboratory results—for more precise assessment of kidney function—are not available in administrative claims. (Figure S2). Among other safety events assessed, dizziness/drowsiness had the next highest incidence rate (69.2 and 62.7) per 1000 years at risk, followed by nausea/vomiting (58.4 and 45.8), diarrhea (36.8 and 35.6), constipation (38.6 and 28.0), skin rash (26.8 and 23.0), and dry mouth (2.8 and 1.2) for the TC and spironolactone cohorts respectively. There were no significant differences in the incidence rates or cumulative incidence for patients newly initiating TC compared with spironolactone after adjusting for differences in baseline characteristics using IPTW for any of these events (Table 3, Figure 2, Figure S2).

## DISCUSSION

In this retrospective longitudinal cohort study using a large sample of recent USA real-world data, the incidence of cardiovascular outcomes and safety events was compared in patients with resistant hypertension without cardiovascular disease history initiating TC versus spironolactone as fourth-line antihypertensive treatment. Results showed that patients with resistant hypertension have similar and low rates of MACE outcomes following initiation of treatment with TC or spironolactone, yet TC appeared to be a safer choice.

It is crucial to have evidence on treatment options for resistant hypertension in terms of cardiovascular protection, rather than only BP lowering effects. Thus, the most important finding of this study is that incidence of MACE was similar with TC and spironolactone among patients with resistant hypertension and no established cardiovascular disease. To the best of our knowledge, a direct comparison of outcomes with these drugs has not been reported in patients with resistant hypertension, either from clinical trials or real-world data analyses.^18^

Another clinical relevant and highly significant finding of this analysis was the lower risk of hyperkalemia among TC users (weighted rate ratio, 0.48; p<0.001), consistent with the known elevated risk with spironolactone.^37^ Using the cumulative incidence after IPTW, among those who received TC, the estimated number needed to harm in 1 year is 58 patients for hyperkalemia if switched to spironolactone. The algorithm used to identify patients with resistant hypertension in the current study, applying statistical weighting methodology and with variable follow-up, was comparable with prior research conducted by Desai et al, which evaluated outcomes among patients initiating aldosterone antagonists versus beta-blockers using administrative claims data^6^. Consistent with the prior analysis, utilization of aldosterone antagonists (spironolactone) was associated with substantially increased risk of hyperkalemia versus the comparator fourth-line agent^10^. It is important to note that in this cohort the doctors certainly avoided prescribing spironolactone in patients with higher risk of hyperkalemia (e.g., those with higher baseline potassium and/or lower GFR), implying that the incidence of hyperkalemia would probably be higher if spironolactone was randomly administered to the participants of this cohort.

The European Society for Hypertension (ESH) guideline notes that clonidine produced a BP-lowering effect similar to that of spironolactone based the REHOT trial,^18^ while acknowledging that this evidence is limited to short-term BP effects rather than cardiovascular outcomes.^38^ This real-world analysis adds an important, complementary dimension. While spironolactone may represent a ‘preferred’ choice as fourth-drug treatment on BP-lowering grounds these data,^17^ combined with REHOT trial,^18^ support that TC can be a “near-peer” option, and a safer one, particularly where spironolactone use is constrained. In addition, the one-weekly administration of TC, may improve long-term adherence in patients with resistant hypertension who usually require complex regimens, and is associated with fewer adverse effects than the oral formulation.^20–22^ Given that spironolactone is currently recommended as a preferred fourth drug in resistant hypertension,^13, 16^ evidence suggesting oral clonidine has a comparable BP lowering effect^17, 18^ together with the present real-world evidence that the transdermal administration of clonidine can achieve comparable cardiovascular outcomes with less hyperkalemia, a practical positioning approach would be placing TC as a closer alternative to spironolactone in cases where: i) hyperkalemia risk is a decisive factor (e.g. patients with reduced estimated glomerular filtration rate, borderline potassium levels or prior hyperkalemia on mineralocorticoid receptor antagonists, or patients on high-risk concomitant renin-angiotensin-aldosterone system inhibitor regimens), ii) treatment adherence is a critical factor (e.g. complex drug regimens, high pill burden), where once-weekly administration of TC may improve long-term adherence, and iii) sympathetic-drive phenotypes (e.g. patients with heightened sympathetic tone contexts, as sleep apnea, CKD, diabetes) where central adrenergic blockade is pharmacologically aligned. The ESH guidelines present a list of conditions in which blocking the sympathetic nervous system with beta-blockers can be beneficial in patients with hypertension, and several of them also apply to central sympathetic system blockade with clonidine.^38^ Moreover, in patients in whom close follow-up to monitor serum potassium is challenging due to healthcare or patient-related factors, avoiding spironolactone is reasonable.

Findings should be interpreted within the context of several study limitations. First, while IPTW was used to balance the cohorts on all measurable characteristics, the potential for residual confounding remains. Administrative claims do not have clinical detail on BP measurements, body mass index, lifestyle factors, or results from laboratory tests all of which may influence the selection of TC or spironolactone. In addition, although resistant hypertension was identified using a rigorous and previously published algorithm, there is the potential for misclassification as BP values and measurement protocol were not available. Therefore, it was not possible to quantify the proportion of our cohort participants who had “apparent” rather than “true” resistant hypertension. Last, this study was limited to individuals without prior history of CVD and with commercial health coverage or private Medicare coverage. Consequently, results of this analysis may not be generalizable to patients with prior CVD history, or with other health insurance or without insurance coverage.

In conclusion, in this real-world USA cohort of patients with resistant hypertension without established CVD, add-on therapy with TC demonstrated similar cardiovascular event risk to spironolactone. However, the risk of hyperkalemia was considerably lower with TC, consistent with known pharmacologic mechanisms. These findings, combined with the pharmacologic characteristics of TC allowing once-per-week administration, support the reconsideration of TC by hypertension guidelines as a useful alternative to spironolactone in patients with resistant hypertension, particularly those at increased risk for hyperkalemia, issues with treatment adherence or regular monitoring, or other medical conditions. Prospective randomized controlled studies with outcome endpoints are needed to confirm these findings.

## NOVELTY AND RELEVANCE

### What Is New?

- This is the first study that compares transdermal clonidine (TC) versus spironolactone in patients with resistant hypertension in terms of cardiovascular outcomes and safety events utilizing large real-world data in the US.

### What Is Relevant?

- Patients with resistant hypertension had similar incidence of major cardiovascular outcomes with add-on treatment with TC or spironolactone, yet significantly higher risk of hyperkalemia with the latter.

### Clinical Implications?

- These findings, combined with the unique pharmacologic characteristics of TC offering once-per-week administration, support the reconsideration of TC by the hypertension guidelines as a useful alternative to spironolactone in patients with resistant hypertension, particularly those at increased risk for hyperkalemia, or other issues related to spironolactone use.

## Data Availability

The data that support the findings of this study are available from Merative. Restrictions apply to the availability of these data, which were used under license for this study.

## ACKNOWLEDGMENTS

Programming services and analyses were supported by Helen Varker from Merative. Statistical analysis services were provided Ryan Ross from Merative. The views expressed in this article are the authors’ own and not an official position of their institutions or funder.

## ABBREVIATIONS

ACEI: Angiotensin-Converting Enzyme Inhibitors
AHR: Adjusted Hazard Ratio
ARB: Angiotensin Receptor Blockers
BP: Blood pressure
CCB: Calcium Channel Blockers
CCI: Charlson Comorbidity Index
CPI: Consumer Price Index
CPT: Current Procedural Terminology
CKD: Chronic Kidney Disease
ESH: European Society for Hypertension
ESRD: End-Stage Renal Disease
GFR: Glomerular filtration rate
HCPCS: Healthcare Common Procedure Coding System
ICD: International Classification of Diseases
IPTW: Inverse Probability of Treatment Weighting
MACE: Major Adverse Cardiovascular Events
NDC: National Drug Codes
PDC: Proportion of Days Covered
SD: Standard Deviation
SMD: Standardized Mean Difference
TC: Transdermal Clonidine

